# Urine complement proteins are associated with kidney disease progression of type 2 diabetes in Korean and American cohorts

**DOI:** 10.1101/2024.08.15.24312080

**Authors:** Donghwan Yun, Sohyun Bae, Yuqian Gao, Lauren Lopez, Dohyun Han, Carrie D. Nicora, Tae Youn Kim, Kyung Chul Moon, Dong Ki Kim, Thomas L. Fillmore, Yon Su Kim, Avi Z. Rosenberg, Weijie Wang, Pinaki Sarder, Wei-Jun Qian, Maryam Afkarian, Seung Seok Han

**Author notes:** Co-first authors (Donghwan Yun, Sohyun Bae). ^†^co-corresponding authors. Correspondence: Seung Seok Han, Department of Internal Medicine, Seoul National University College of Medicine 103 Daehak-ro, Jongno-gu, Seoul, 03080, Republic of Korea, Maryam Afkarian, Room 5317, Genome and Biomedical Sciences Facilities, 451 Health Sciences Drive, University of California, Davis, CA 95616, USA, Tet.

## Abstract

**Background:** Mechanisms of progression of diabetic kidney disease (DKD) are not completely understood. This study uses untargeted and targeted mass spectrometry-based proteomics in two independent cohorts on two continents to decipher the mechanisms of DKD in patients with type 2 diabetes.

**Methods:** We conducted untargeted mass spectrometry on urine samples collected at the time of kidney biopsy from Korean patients with type 2 diabetes and biopsy-proven diabetic nephropathy at Seoul National University Hospital (SNUH-DN cohort; n = 64). These findings were validated using targeted mass spectrometry in urine samples from a Chronic Renal Insufficiency Cohort subgroup with type 2 diabetes and DKD (CRIC-T2D; n = 282). Urinary biomarkers/pathways associated with kidney disease progression (doubling of serum creatinine, ≥50% decrease in estimated glomerular filtration rates, or the development of end-stage kidney disease) were identified.

**Results:** SNUH-DN patients had an estimated glomerular filtration rate (eGFR) of 55 mL/min/1.73 m^2^ (interquartile range [IQR], 44–75) and random urine protein-to-creatinine ratio of 3.1 g/g (IQR, 1.7–7.0). Urine proteins clustered into two groups, with cluster 2 having a 4.6-fold greater hazard (95% confidence interval [CI], 1.9–11.5) of disease progression than cluster 1 in multivariable-adjusted, time-to-event analyses. Proteins in cluster 2 mapped to 10 pathways, four of the top five of which were complement or complement-related. A high complement score, constructed from urine complement protein abundance, was strongly correlated to 4 of 5 histopathologic DN features and was associated with a 2.4-fold greater hazard (95% CI, 1.0–5.4) of disease progression than a low complement score. Targeted mass spectrometry of the CRIC-T2D participants, who had an eGFR of 42 mL/min/1.73 m^2^ (IQR, 37–49) and 24-hr urine protein of 0.48 g (IQR, 0.10–1.87), showed that the complement score similarly segregated them into rapid and slow DKD progression groups. In both cohorts, the complement score had a linear association with disease progression.

**Conclusions:** Urinary proteomic profiling confirms the association between the complement pathway and rapid DKD progression in two independent cohorts. These results suggest a need to further investigate complement pathway inhibition as a novel treatment for DKD.

## Introduction

Diabetic kidney disease (DKD) is a major complication of diabetes, significantly contributing to morbidity and mortality worldwide ^1, 2^. As the leading cause of end-stage kidney disease (ESKD) globally, DKD accounts for about half of all ESKD cases, thereby posing a significant burden on healthcare systems worldwide ^3^. Despite recent advances in diabetes management, the mechanisms underlying progression (and their respective biomarkers) in human DKD remain incompletely understood. Clinically, this translates into highly variable rates of disease progression, with some patients experiencing rapid progression leading to ESKD ^4, 5^. This variability presents substantial challenges in clinical practice, as it is currently impossible to identify and intervene in patients who are rapid progressors.

Although extensive research is being conducted, the pathophysiological mechanisms responsible for the DKD progression still need to be completely understood. Traditional risk factors, such as hyperglycemia, hypertension, and diabetic duration, are recognized ^6, 7^, but they only partially explain the inter-individual differences in disease progression. Prior studies have explored various biomarkers, including cytokines and kidney injury-relevant molecules ^8–10^, but a comprehensive understanding that integrates these factors with clinical outcomes remains elusive. This knowledge gap hinders the development of precise predictive models and targeted interventions that are essential for preventing rapid progression in DKD patients. Therapeutic strategies such as renin-angiotensin-system inhibitors, sodium-glucose cotransporter-2 inhibitors, and mineralocorticoid receptor antagonists are employed to mitigate DKD progression ^11–13^. However, despite these advances in therapy, it is evident that more targeted therapies are needed to improve DKD outcomes.

In response to these challenges, -omics technologies, including genomics, transcriptomics, metabolomics, and proteomics, emerge as powerful tools for unraveling the complex pathobiology of DKD. Recent advances in urinary proteomics have significantly enhanced the ability to predict prognosis and analyze subtypes in DKD by revealing the molecular dynamics of kidney injury ^14, 15^. This approach allows for the development of personalized treatment plans based on specific molecular profiles. We used comparative proteomic analyses across two independent cohorts (South Korea and the US) to identify the pathways associated with rapid DKD progression in type 2 diabetes. Utilizing both untargeted and targeted urinary proteomics, we found that differential abundance of complement proteins in urine predicted rapid DKD progression. This data highlights the importance of the complement pathway as a promising source of novel biomarkers for identifying rapid progressors and as a therapeutic target for novel DKD interventions.

## Methods and Materials

### Study patients

From March 2011 to June 2021, 93 patients with type 2 diabetes and biopsy-proven diabetic nephropathy were recruited at Seoul National University Hospital (SNUH), referred to as the SNUH-DN cohort. Of this group, 29 were excluded due to an estimated glomerular filtration rate (eGFR) <30 mL/min/1.73 m^2^ (n = 25), combined type 1 and type 2 diabetes (n = 2), the presence of concomitant nondiabetic glomerular disease (n = 1), or the absence of glomeruli in biopsied kidney tissues (n = 1). The remaining 64 patients were included in the final analyses as the SNUH-DN cohort after conducting untargeted proteomics.

Additionally, this study incorporated a subset of patients with type 2 diabetes from the Chronic Renal Insufficiency Cohort (CRIC) study, termed CRIC-T2D, consisting of American patients with risk factors for cardiovascular disease, progression of chronic kidney disease, and mortality. A total of 3,939 patients aged 21–74 with an eGFR of 20–70 mL/min/1.73 m^2^ were enrolled at seven clinical centers throughout the US from June 2003 to December 2008. CRIC over-sampled participants with diabetes, who constitute nearly half of the CRIC cohort. This study excluded participants with monogenic renal disease, liver cirrhosis, class III or IV heart failure, human immunodeficiency virus, cancer, autoimmune disease, polycystic kidney disease, pregnant women, recipients of organ or bone marrow transplants, and those receiving immunotherapy for primary renal disease or systemic vasculitis within the six months preceding recruitment or systemic chemotherapy.

### Ethical considerations

This study was conducted in accordance with the ethical principles outlined in the Declaration of Helsinki and was approved by the institutional review boards (IRBs) of all participating institutions. For the SNUH-DN cohort, ethical approval was granted by the IRB at SNUH (no. H-2403-066-1519). The CRIC study was approved by the IRBs at all participating institutions and was conducted in accordance with the Declaration of Helsinki ^16, 17^. Urine samples and data for the present study were obtained from the CRIC subset housed at the NIDDK Central Repository (R01 5R01DK104706). The use of de-identified CRIC samples was approved by the IRB at the University of California, Davis. The use of de-identified samples and data from the CRIC-T2D subgroup from the NIDDK Central Repository was approved by the Human Subjects Division, University of California, Davis. All participants provided written informed consent, having been fully informed of the study purpose, the procedures involved, potential risks, and their rights as study participants, including the right to withdraw at any point without consequence. The confidentiality of patient data was rigorously maintained throughout the study. All personal identifiers for SNUH-DN patients were removed, and data was stored securely to prevent unauthorized access. CRIC-T2D data and samples were de-identified at the NIDDK Central Repository.

### Study outcomes

The primary outcome was the progression of DKD (named kidney disease progression) based on any of the following events: a doubling of baseline serum creatinine, a decline greater than 50% in the eGFR, or the onset of ESKD. The eGFR was calculated using the Chronic Kidney Disease Epidemiology Collaboration equation ^18^. ESKD was specifically defined as the need for kidney replacement therapy, including hemodialysis, peritoneal dialysis, or kidney transplantation.

### Study variables

We collected comprehensive patient data encompassing demographic characteristics, such as age, sex, race, body mass index, diabetic duration, and comorbidities of hypertension, ischemic heart disease, cerebrovascular disease, retinopathy, and neuropathy. Laboratory findings included hemoglobin A1c, eGFR, and random urine protein-to-creatinine ratio (PCR) for the SNUH-DN cohort, while the CRIC-T2D cohort included the 24-hour urine protein amount.

We employed a detailed histopathologic classification system to evaluate kidney biopsies from patients diagnosed with diabetic nephropathy in the SNUH-DN cohort. This classification system follows the criteria described in the reference paper ^19^. Each biopsy was required to contain at least ten glomeruli for accurate assessment, excluding any incomplete glomeruli along the biopsy edges. The glomerular classification was segmented into five categories: class I, isolated glomerular basement membrane thickening; class IIA, mild mesangial expansion; class IIB, severe mesangial expansion; class III, nodular sclerosis (Kimmelstiel-Wilson lesions); and class IV, advanced diabetic glomerulosclerosis with more than 50% global glomerulosclerosis. The assessment of interstitial fibrosis and tubular atrophy (IFTA), interstitial inflammation, arterial hyalinosis, and arteriosclerosis was also quantified using a four-point or three-point scale according to the criteria.

### Urine sample collection

In the SNUH-DN cohort, urine samples were collected at the time of kidney biopsy. Patients were instructed to provide a midstream clean-catch urine sample, which was immediately processed to minimize protein degradation. Upon collection, urine samples were centrifuged at 3000 rpm for 10 minutes to remove cellular debris and then aliquoted into cryovials. The aliquots were promptly frozen and stored at –80°C to preserve the integrity of the proteins until preparation for proteomics.

In the CRIC-T2D cohort, 24-hour urine samples were collected at home by study participants during their CRIC visit number 3. Participants were instructed to keep the samples at 4°C during collection. Collections that were less than 500 ml or collected for <22 hours or >26 hours were repeated. Accepted 24-hour collections were thoroughly mixed, and three 10 ml aliquots were obtained. The aliquots were shipped to the CRIC central laboratory on ice packs, where they were further aliquoted and stored at –80°C. A portion of these samples was shipped to the NIDDK Central Repository when they were sent to a study author (MA). An aliquot of these samples was then shipped to the Pacific Northwest National Laboratory on dry ice and were prepared for targeted proteomics.

### Untargeted proteomics

#### Sample preparation

A volume of 1-2 ml of urine from each participant was concentrated down to 250 µl using a centrifugal filter with a molecular weight cutoff of 3 kDa (Millipore, Billerica, MA). Protein concentrations were then determined using the Bradford method, employing a commercial kit (Bio-Rad, Hercules, CA). For the analysis, 50 µg of protein from each sample was precipitated using ice-cold acetone and added at five times the protein volume. The precipitate was redissolved in 50 µl of SDT buffer (comprising 2% SDS, 0.1 M dithiothreitol, in 0.1 M Tris HCl, pH 8.0), and subjected to heating at 95°C to denature the proteins. Following denaturation, proteins were enzymatically digested utilizing a filter-aided sample preparation approach, adapted from previously established methods with some modifications ^20^. Specifically, proteins were transferred to an Amicon 30K filter (Millipore, Billerica, MA) and washed multiple times with UA solution (8 M urea in 0.1M Tris-HCl, pH 8.5) through centrifugation. After three washes, cysteines were alkylated using 0.05 M iodoacetamide in the UA solution for 30 minutes at room temperature in the dark. Subsequent buffer exchanges to 40 mM ammonium bicarbonate were performed twice to prepare for enzymatic digestion. Overnight digestion was carried out at 37°C using trypsin/LysC with a ratio of enzyme to substrate of 1:100. The peptides produced were then collected in new tubes through additional centrifugation, and an extra elution step was performed using 40 mM ammonium bicarbonate mixed with 0.5 M sodium chloride. The eluted peptides were purified and fractionated using custom-made styrene-divinylbenzene reversed-phase sulfonate-StageTips, employing a gradient of acetonitrile (40%, 60%, and 80%) in 1% ammonium hydroxide ^21, 22^. Following fractionation, peptides were vacuum-dried and stored at – 80°C until further analysis.

### Construction of a peptide spectral library

To create a peptide spectral library for using match between runs (MBRs) algorithm of maxquant, a pooling of urine proteins was performed ^23^. We combined equal sub-aliquots of proteins, each containing 5 µg, to a total of 100 µg. This pooled sample was then subjected to a two-step filter-aided digestion process, consistent with the methods previously outlined ^21, 24^. Following digestion, peptides were purified using Oasis HLB solid-phase extraction cartridges to ensure their readiness for high-performance liquid chromatography. For the library construction, we utilized 100 µg of these clean peptides and processed them through an Agilent 1260 bioinert HPLC system (Agilent, Santa Clara, CA) equipped with a standard analytical column (4.6 × 250 mm; 5-µm particle size). The peptides were fractionated using a high-pH reversed-phase method, employing a flow rate of 0.8 ml/min across a 60-minute gradient. Solvents included 15 mM ammonium hydroxide in water (solvent A) and 15 mM ammonium hydroxide in 90% acetonitrile (solvent B). We collected 96 separate fractions at 30-second intervals throughout a 48-minute period, which were then concatenated into 24 fractions in step-wise manner. Early-, middle-, and late-eluting peptides were pooled by mixing every 24^th^ original fraction for the proteome (e.g., combining fractions 1, 25, 49, and so on) ^25^. These concatenated fractions were subsequently dried using a vacuum centrifuge and stored at –80°C, ready for analysis by liquid chromatography-tandem mass spectrometry (LC-MS/MS).

### LC-MS/MS analytical procedure

LC-MS/MS analysis was conducted on a Q-Exactive Plus Quadrupole Orbitrap mass spectrometer (Thermo Fisher Scientific, Waltham, MA), which was interfaced with an Ultimate 3000 RSLC nano system (Dionex, Sunnyvale, CA) featuring a nanoelectrospray ion source. The process incorporated slight modifications from previously established protocols ^20, 22^. Individual peptide fractions from urine samples underwent separation using a dual-column arrangement consisting of a C18 trap column (300 µm internal diameter × 0.5 cm, with 3 µm particles and 100 Å pore size) and a C18 analytical column (50 µm internal diameter × 50 cm, with 1.9 µm particles and 100 Å pore size). Prior to injection, dried peptide fractions were dissolved in solvent A (2% acetonitrile and 0.1% formic acid). The nano-LC system loaded the samples and applied a chromatographic gradient over 90 minutes, increasing from 8% to 30% of solvent B (100% acetonitrile with 0.1% formic acid). The setup operated under a spray voltage of 2.0 kV in positive ion mode, and the capillary heater was maintained at 320°C. Mass spectrometric data were collected in a data-dependent acquisition format, prioritizing the top 15 most abundant precursor ions. The Orbitrap detector analyzed precursor ions across a mass range from 300 to 1650 m/z, achieving a resolution of 70,000 at m/z 200. Fragmentation was performed via higher-energy collisional dissociation (HCD) at a resolution of 17,500 for m/z 200, using a normalized collision energy setting of 28. The system’s ion injection times were set to a maximum of 20 ms for survey scans and 120 ms for MS/MS scans.

### Label-free quantitative data analysis

For the analysis of mass spectrometric data, we employed MaxQuant software (version 1.6.1.0) for label-free quantification ^26^. MS/MS spectra were meticulously matched against the Human UniProt protein sequence database (as of December 2014, containing 88,657 entries) via the Andromeda search engine ^27^. The analysis parameters were carefully selected, with a precursor ion tolerance set at 6 ppm for total protein analysis and an MS/MS ion tolerance of 20 ppm. We designated carbamido-methylation of cysteine as a fixed modification, while N-acetylation of proteins and methionine oxidation was considered variable modifications. The digestion parameter was configured for full tryptic digestion with allowances for up to two missed cleavages per peptide, which had to be at least six amino acids long. We rigorously maintained a false discovery rate (FDR) of 1% across peptide, protein, and modification levels to ensure high data integrity. Additionally, to enhance quantification accuracy across samples, we utilized a strategy of matching between runs anchored by the peptide library constructed from pooled urine samples. The Intensity Based Absolute Quantification (iBAQ) algorithm, a component of the MaxQuant suite, facilitated the estimation of protein abundances ^28^. In iBAQ, raw intensities were normalized by the count of theoretical peptides, rendering them reflective of the proteins’ molar quantities.

### Targeted proteomics

#### Selected reaction monitoring (SRM)

Assay development was conducted as described in detail previously ^29^. Briefly, final surrogate peptides were selected based on the uniqueness of their occurrence in proteins of interest and their presence and chromatographic behavior in shotgun MS/MS data in our prior urine proteomics data repository (Supplementary Table 1). Crude synthetic heavy isotope-labeled versions of the selected peptides were used to generate MS/MS data in an Orbitrap HCD. This data was used to identify the best transitions and optimal collision energy for each transition.

### Sample processing and digestion

Urine samples were processed and underwent LC-MS/MS, as described previously ^29^. Briefly, urine samples were thawed on ice, run through a 10 kDa prewashed (50 mM AMBIC, i.e., NH_4_HCO_3_, pH 8.0) Amicon ultrafiltration column, and underwent buffer exchange with AMBIC twice. The final urine protein concentration was determined using the BCA protein assay. Urea was added to the urine sample to a final concentration of 8 M, followed by reduction with 500mM DTT to a final concentration of 10 mM, brief sonication, and incubation at 37°C for 1 hour while shaking. Alkylation was performed by adding IAA to a final concentration of 40 mM and incubation for one hour at 37°C in the dark while shaking. The sample was then diluted ten-fold with digestion buffer, and trypsin was added to a protein/trypsin ratio of 50:1 (w/w), followed by incubation at 37°C for 3 hours. Digestion was stopped by adding TFA to a final concentration of 0.1%. The tryptic digest was separated using SPE C18 columns, preconditioned using methanol and then SPE conditioning buffer, and washed with SPE washing buffer. Peptides were eluted from the SPE C18 columns with 1ml SPE elution buffer and, dried under a reduced vacuum using a speed vac and redissolved in MS-grade water. The peptide concentration was determined using the BCA protein assay, and peptide samples were stored at –80°C until further use. Heavy isotope-labeled peptide standards were mixed in a single 1 ml stock solution, having been reconstituted at 1000 fmol/ml per peptide in 0.1% TFA in water. The digested peptide mix and the heavy peptide stock were combined while shaking and advanced to LC-SRM analysis.

### LC-SRM analysis and quantification

The urine sample/heavy peptide mixes were analyzed using the final transition list, derived from the orbitrap HCD MS/MS, and optimized in one typical urine sample. LC-SRM was performed using a nanoACQUITY UPLC system coupled online to a TSQ Vantage triple quadrupole mass spectrometer. The peak area ratios (PAR) of endogenous transitions and heavy internal standard transitions represent the molar ratios between the amounts of endogenous and heavy internal standard peptides. The PARs were calculated using Skyline software ^30^. The data was imported into a Skyline file, peak boundaries for each peptide were manually verified for each dataset, and detection and optimal transition (for best peak area ratio, PAR) were confirmed for each peptide. Protein concentration was calculated using the equation below.

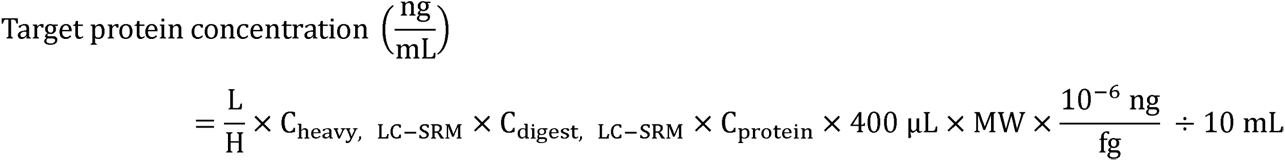

where L/H is PAR of endogenous (L) to heavy isotope-labeled internal standard peptide (H), C_heavy_, LC-SRM is the molar concentration of heavy internal standard peptides (fmol/μl) in the final LC-SRM solutions; MW is the molecular weight of targeted protein (Da or g/mol); C_digest, LC-SRM_ is the mass concentration of total protein digest (μg/μl) in the final LC-SRM solutions; and C_protein_ is mass concentration of total protein (μg/μl) in the 400 μl concentrated retentate from 10 ml of original urine.

### Proteomics data processing and analysis

The proteomic data obtained using label-free quantification of urine proteins were initially processed using the *Seurat* package (version 5.0.3) ^31^. These values were normalized for urine dilution by indexing (division) by urine creatinine, and then log2 transformed for further normalization using the *NormalizeData* function from the *Seurat* package. Finally, for each identified protein, the normalized data was scaled using the *ScaleData* function from the Seurat. Unsupervised clustering techniques were utilized to group proteins based on their expression patterns, and the *FindNeighbor* function and *RunUmap* function were used to create cluster groups and to draw UMAP, respectively. Gene names related to each protein were identified using the *UniProt* accession numbers ^32^. Differentially expressed proteins between rapid and slow progressors were identified (*FindMarkers* function). Using the *UniProt* database, proteins with well-documented gene associations were selected for pathway analysis, and differentially represented pathways were identified using *WikiPathways* ^33^.

### Complement scores

The complement score was devised to evaluate the prevalence of complement pathway proteins in urinary proteomics data. We identified the proteins involved in the complement pathway by referencing the Kyoto Encyclopedia of Genes and Genomes database ^34^. The list of complement proteins used for scoring is presented in Supplementary Table 2. The average protein expression of denoted genes was calculated based on the assumption of previous studies ^35–37^. Additionally, we formalized the distinction between activators and inhibitors. For each patient, this score was calculated by taking the mean of the normalized abundance of identified complement proteins, adjusted for their functional roles: inhibitory proteins were assigned a modifier of –1, and activating proteins a modifier of 1. The formula used was:

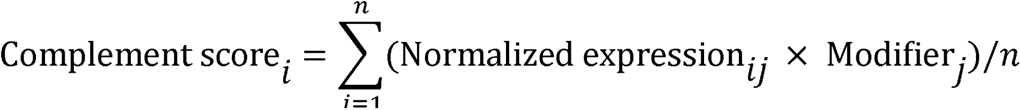

where *i* denotes the sample number, and *j* is the individual complement protein. The continuous variable complement score was binarized into low vs. high complement score bins by the median value.

### Statistical analysis

Descriptive statistical analyses were conducted on patient information. For continuous variables with a normal distribution, data are presented as the mean ± standard deviation, and for those without a normal distribution, as medians with interquartile ranges (IQR). The Kolmogorov-Smirnov test was used to assess the normality of the data distribution. Categorical variables are shown as percentages. Comparison of categorical variables was performed using either the chi-square test or Fisher’s exact test, while continuous variables were compared using either Student’s t-test or the Mann-Whitney U test, depending on their distribution. When comparing multiple groups for continuous variables, analysis of variance (ANOVA) was used, with the groups treated as ordinal variables. Pairwise comparisons were conducted with Bonferroni correction.

Survival analysis was employed to investigate the association between variables and the risk of kidney disease progression. The hazard ratio (HR) and corresponding 95% confidence intervals were calculated using the Cox proportional hazards regression model. Variables included in the model were selected based on clinical relevance, such as age, sex, body mass index, diabetic duration, and baseline kidney function. Adjusted survival curves were then generated to visualize the differences in adjusted cumulative survival between the two groups using the *adjustedCurves* package ^38^. Additionally, penalized spline regression was utilized to further elucidate the relationship between complement scores and kidney disease progression. All statistical analyses were conducted using the R program (version 4.2.2). A two-sided *P* value of less than 0.05 was considered statistically significant.

## Results

### Baseline characteristics in the SNUH-DN cohort

This cohort had a mean age of 53.3 ± 11.1 years, with 71.9% male subjects. Hypertension was present in 92.2% of the subjects, and the median duration of diabetes was 10 years (IQR, 4– 14 years). Diabetic retinopathy and neuropathy were observed in 54.7% and 29.7% of all patients, respectively. The median hemoglobin A1c was 7.2% (IQR, 6.6–8.4), while the median PCR and eGFR values were 3.1 g/g (IQR, 1.7–7.0) and 55 mL/min/1.73 m² (IQR 44–75), respectively. Details of histopathologic features are provided in Table 1.

**Table 1.** Baseline characteristics of the SNUH-DN cohort.

| Variables | Total<br>(n = 64) | Cluster 1<br>(n = 40) | Cluster 2<br>(n = 24) | *P* |
| --- | --- | --- | --- | --- |
| Age (years) | 53.3 ± 11.1 | 52.1 ± 10.9 | 55.2 ± 11.5 | 0.282 |
| Male (%) | 71.9 | 75.0 | 66.7 | 0.667 |
| Body mass index (kg/m²) | 25.0 ± 3.3 | 25.2 ± 3.1 | 24.7 ± 3.5 | 0.594 |
| Hypertension (%) | 92.2 | 95.0 | 87.5 | 0.355 |
| Diabetic duration (years) | 10 (4–14) | 8 (2–12) | 11 (4–20) | 0.278 |
| Diabetic retinopathy (%) | 54.7 | 52.5 | 58.3 | 0.846 |
| Diabetic neuropathy (%) | 29.7 | 30.0 | 29.2 | 1.000 |
| Ischemic heart disease (%) | 7.8 | 12.5 | 0 | 0.148 |
| Cerebrovascular disease (%) | 4.7 | 2.5 | 8.3 | 0.551 |
| Hemoglobin A1c (%) | 7.2 (6.6–8.4) | 7.2 (6.6–7.6) | 7.4 (6.8–8.9) | 0.178 |
| PCR (g/g) | 3.1 (1.7–7.0) | 2.2 (1.2–3.6) | 7.1 (4.9–9.5) | <0.001 |
| eGFR (mL/min/1.73 m²) | 56 (44–75) | 61.2 (48–85) | 43 (33–60) | <0.001 |
| Glomerular classification (%) |  |  |  |  |
| I | 14.1 | 17.5 | 8.3 | 0.003 |
| IIA | 12.5 | 17.5 | 4.2 |  |
| IIB | 32.8 | 42.5 | 16.7 |  |
| III | 12.5 | 10.0 | 16.7 |  |
| IV | 28.1 | 12.5 | 54.2 |  |
| Interstitial fibrosis and Tubular atrophy (%) |  |  |  |  |
| 0 | 4.7 | 7.5 | 0.0 | <0.001 |
| 1 | 35.9 | 47.5 | 16.7 |  |
| 2 | 46.9 | 45.0 | 50.0 |  |
| 3 | 12.5 | 0.0 | 33.3 |  |
| Interstitial Inflammation (%) |  |  |  |  |
| 0 | 10.9 | 17.5 | 0.0 | 0.044 |
| 1 | 84.4 | 80.0 | 91.7 |  |
| 2 | 4.7 | 2.5 | 8.3 |  |
| Arteriolar hyalinosis (%) |  |  |  |  |
| 0 | 25.0 | 35.0 | 8.3 | 0.052 |
| 1 | 28.1 | 22.5 | 37.5 |  |
| 2 | 46.9 | 42.5 | 54.2 |  |
| Arteriosclerosis (%) |  |  |  |  |
| 0 | 25.0 | 27.5 | 20.8 | 0.674 |
| 1 | 35.9 | 37.5 | 33.3 |  |
| 2 | 39.1 | 35.0 | 45.8 |  |
| Follow-up time (years) | 6.2 (2.4–7.8) | 7.1 (4.9–8.4) | 2.7 (1.1–5.1) | <0.001 |

PCR, random urine protein-to-creatinine ratio; eGFR, estimated glomerular filtration rate.

### Proteomic clustering in the SNUH-DN cohort

Unsupervised clustering of the iBAQ-normalized urine protein abundances from the SNUH-DN cohort identified two distinct clusters, designated as clusters 1 and 2 (Figure 1A). Heatmap of iBAQ-normalized urine proteins that were abundant and scarce in clusters 0 and 1 are illustrated in Supplementary Figures 1A and B, respectively. These data showed that of the 1,877 identified proteins, 71 and 85 were significantly abundant and scarce in cluster 2, compared to cluster 1, after the Bonferroni adjustment, respectively. Many of the abundant proteins mapped to the complement pathway, either complement activators such as complement factor D (CFD), component 2 (C2), component 8 beta chain (C8B), component 6 (C6), and component 5 (C5) ^39^, or complement inhibitors such as complement factor H (CFH) and serpin family G member 1 (SERPING1) ^40, 41^. Conversely, among the scarce proteins, CD55 and CD59 act as complex inhibitors ^42^, while the protein C receptor (PROCR) functions indirectly as a complement inhibitor ^43^.

**Figure 1.**
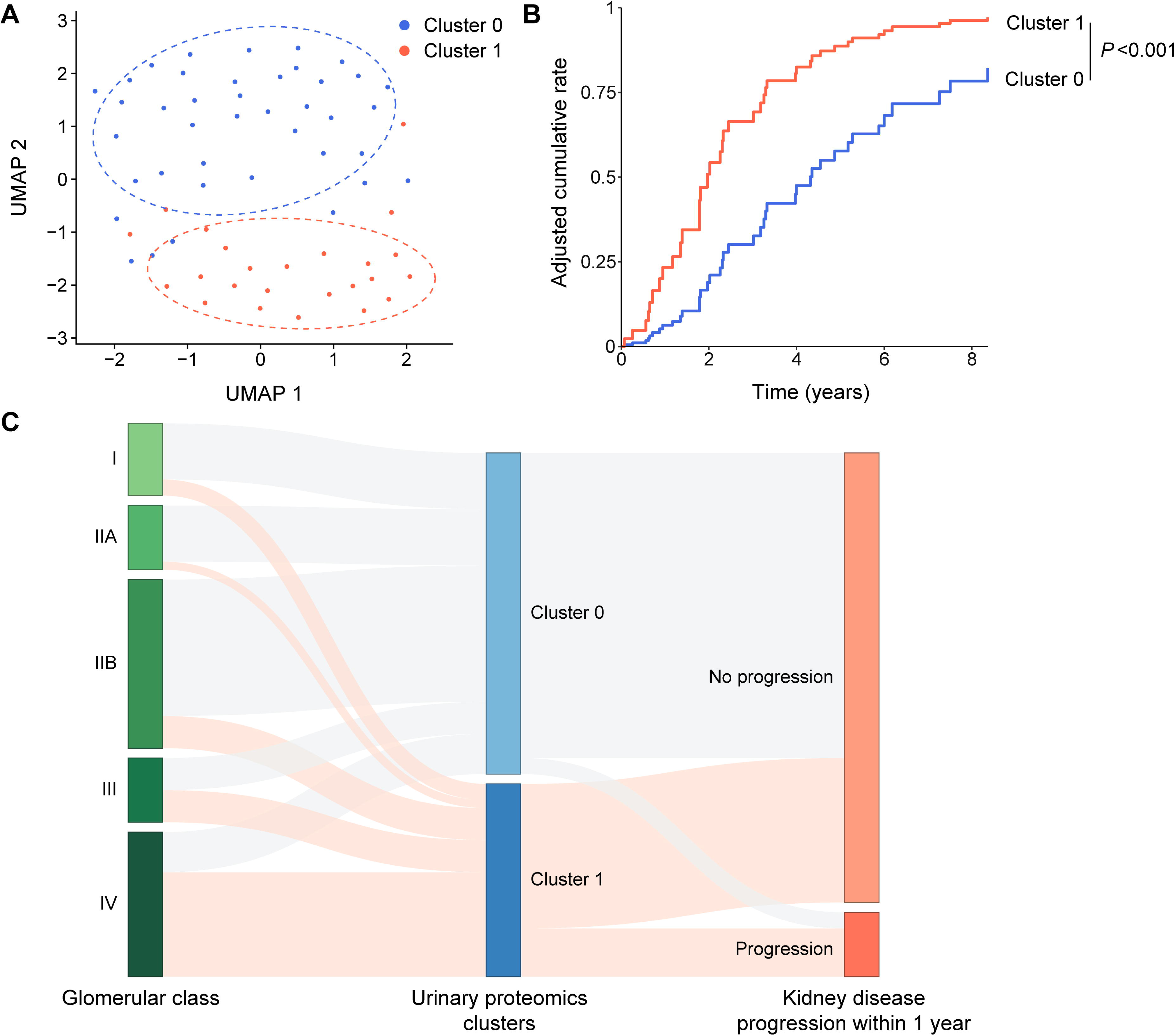
Unsupervised clustering based on untargeted proteomics in the SNUH-DN cohort. **A.** Uniform manifold approximation and projection (UMAP) plot with two distinct clusters (clusters 1 and 2). **B.** Multivariable-adjusted cumulative incidence of kidney disease progression according to clusters, after adjustment for age, sex, body mass index, hypertension, diabetic duration, urine protein-to-creatine ratio, and eGFR. **C.** Sankey plot illustrating the relationships among glomerular classes, clusters, and kidney disease progression within one year.

Kidney disease progression within one year after biopsy occurred in 6 out of 24 subjects (25%) with urine proteins in cluster 2, and 2 out of 40 subjects (5%) in cluster 1. Compared to subjects in cluster 1, those in cluster 2 had a 4.62-fold (95% CI, 1.86–11.50) greater hazard of DKD progression in multivariable time-to-event (Cox proportional hazard) regression, adjusted for age, sex, body mass index, hypertension, diabetic duration, PCR, and eGFR (Table 2). The adjusted cumulative incidence of DKD progression displayed more rapid kidney disease progression in cluster 2 than in cluster 1 (Figure 1B). These results indicate that the unsupervised clustering of urine proteins was able to segregate subjects into two clusters, displaying slower and more rapid DKD progression. Notably, the accelerated DKD progression in cluster 2, compared to cluster 1, was associated with high glomerulosclerosis scores in the paired kidney biopsies from these subjects (Figure 1C).

**Table 2.** Association between cluster and kidney disease progression in the SNUH-DN cohort.

| Variables | Univariable |  | Multivariable* |  |
| --- | --- | --- | --- | --- |
|  | HR (95% CI) | *P* | HR (95% CI) | *P* |
| Cluster 2 (vs. 1) | 4.63 (2.16–9.93) | <0.001 | 4.62 (1.86–11.50) | 0.001 |
| Age (per 1 year) | 1.01 (0.98–1.03) | 0.759 | 0.94 (0.90–0.98) | 0.004 |
| Female (vs. male) | 0.47 (0.20–1.08) | 0.074 | 0.27 (0.10–0.70) | 0.007 |
| Body mass index (per 1 kg/m<sup>2</sup>) | 0.93 (0.83–1.04) | 0.224 | 0.88 (0.78–0.99) | 0.039 |
| Hypertension (vs. none) | 1.14 (0.35–3.74) | 0.827 | 1.03 (0.29–3.68) | 0.964 |
| Diabetic duration (per 1 year) | 1.02 (0.98–1.07) | 0.380 | 1.00 (0.95–1.05) | 0.969 |
| PCR (per 1 g/g) | 1.16 (1.08–1.25) | <0.001 | 1.16 (1.06–1.28) | 0.002 |
| eGFR (per 1 mL/min/1.73 m<sup>2</sup>) | 0.98 (0.96–0.99) | 0.006 | 0.99 (0.97–1.00) | 0.142 |

\*Adjusted for age, sex, body mass index, hypertension, diabetic duration, proteinuria, and estimated glomerular filtration rate.
HR, hazard ratio; CI, confidence interval; PCR, random urine protein-to-creatinine ratio; eGFR, estimated glomerular filtration rate.

### Different enrichment in complement proteins

Pathway analysis identified a notable enrichment of complement and related pathways in cluster 2 (Figure 2A and Supplementary Table 3). The volcano plot of differentially abundant proteins highlighted that complement activators, including CFD, component 3 (C3), C5, C6, and C8B, and some complement inhibitors, including CFH, complement factor I (CFI), and clusterin (CLU), were prominently ranked in cluster 2. Conversely, other complement inhibitors (CD55 and CD59) were scarce in cluster 2 (Figure 2B). Detailed information regarding the proteins related to the complement pathway is included in Supplementary Table 2. The heatmap with the normalized abundance of complement proteins depicted that complement pathway-related proteins were more prominently altered in cluster 2 than in cluster 1 (Figure 2C).

**Figure 2.**
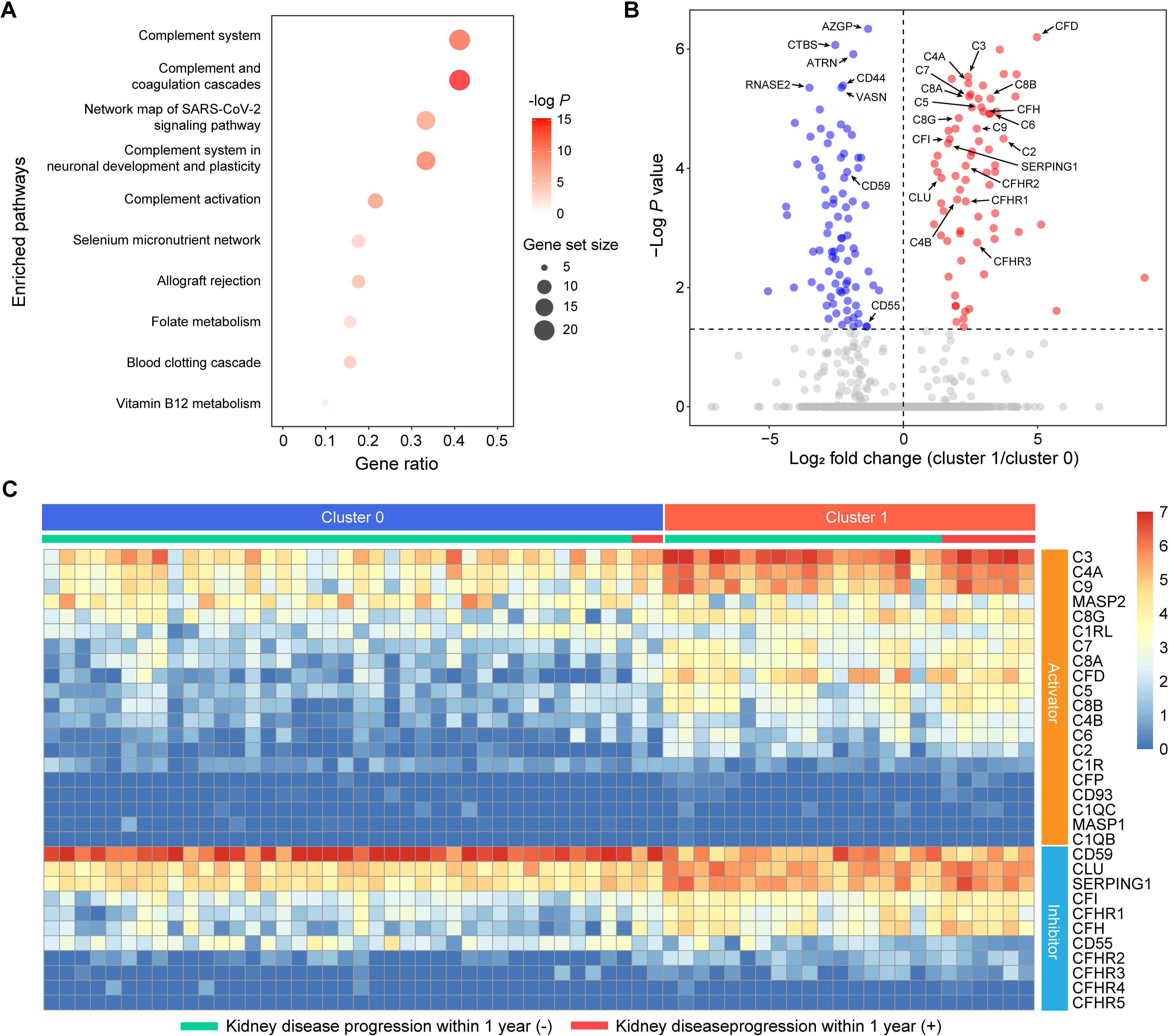
Complement pathway and kidney disease progression in the SNUH-DN cohort. **A.** Pathway analysis of the differentially abundant proteins in cluster 2 compared to cluster 1. **B.** Volcano plot of the differentially abundant proteins in cluster 2 compared to cluster 1. The dashed line denotes the threshold of significance. Red and blue dots represent proteins increased and decreased in cluster 2, respectively, compared to cluster 1. **C.** Heatmap of protein abundance for urinary complement components according to clustering and complement function.

### Complement score and kidney disease progression in the SNUH-DN cohort

To comprehensively assess the effect of complement proteins, we calculated a complement score that considers their expression levels and modifies their contribution by their roles in pathway activation (activators or inhibitors). The overall median complement score for all samples was 0.28 (IQR, 0.08–0.62), with a range from –0.25 to 0.92. Cluster 2 had a median complement score of 0.69 (IQR, 0.51–0.76), while cluster 1 had a median score of 0.14 (IQR, 0.01–0.28) (*P* <0.001) (Figure 3A). Based on the median score of 0.28 from all samples, we divided the patients into high (>0.28) and low (≤0.28) complement score groups (Supplementary Figure 2).

**Figure 3.**
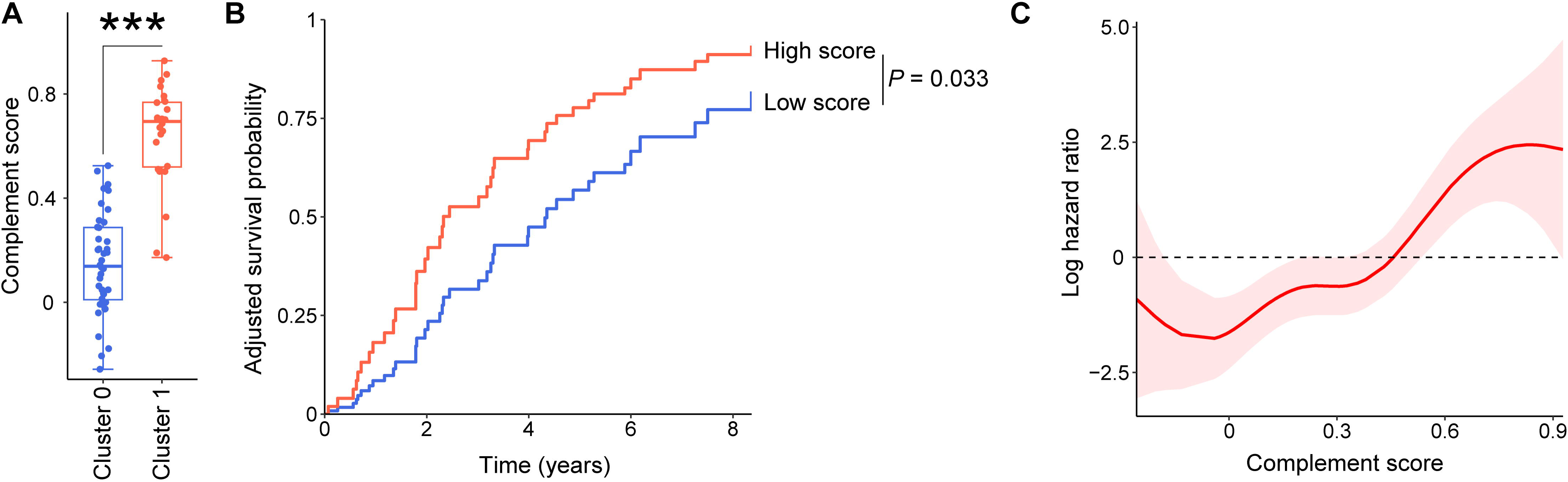
Complement score and kidney disease progression in the SNUH-DN cohort. **A.** Bar plot of complement score in clusters 1 and 2. \*\*\**P* <0.001. **B.** Multivariable adjusted cumulative incidence of kidney disease progression according to complement score groups after adjustment for age, sex, body mass index, hypertension, diabetic duration, urine protein-to-creatinine ratio, and eGFR. **C.** Penalized spline regression according to complement score after adjusting for age, sex, body mass index, hypertension, diabetic duration, urine protein-to-creatinine ratio, and eGFR.

Patients with high complement scores had a 2.41-fold (1.07–5.40) higher risk of kidney disease progression than those with low scores, after adjustment for demographic and clinical variables (Table 3). Multivariable-adjusted cumulative incidence of DKD progression also demonstrated a significant difference between the high and low complement groups (Figure 3B). Notably, in multivariable-adjusted spline regression, the association between the complement score and the hazard of DKD progression was linear (Figure 3C).

**Table 3.** Association between complement score and kidney disease progression in the SNUH-DN cohort.

| Variables | Univariable |  | Multivariable* |  |
| --- | --- | --- | --- | --- |
|  | HR (95% CI) | *P* | HR (95% CI) | *P* |
| High complement score (vs. low) | 2.67 (1.37–5.18) | 0.004 | 2.41 (1.07–5.40) | 0.033 |
| Age (per 1 year) | 1.00 (0.98–1.03) | 0.759 | 0.95 (0.91–0.99) | 0.011 |
| Female (vs. male) | 0.47 (0.20–1.08) | 0.074 | 0.30 (0.11–0.79) | 0.015 |
| Body mass index (per 1 kg/m<sup>2</sup>) | 0.93 (0.83–1.04) | 0.224 | 0.88 (0.78–1.00) | 0.050 |
| Hypertension (vs. none) | 1.14 (0.35–3.74) | 0.827 | 0.89 (0.25–3.11) | 0.852 |
| Diabetic duration (per 1 year) | 1.02 (0.98–1.07) | 0.380 | 1.03 (0.98–1.08) | 0.311 |
| PCR (per 1 g/g) | 1.16 (1.08–1.25) | <0.001 | 1.14 (1.03–1.26) | 0.010 |
| eGFR (per 1 mL/min/1.73 m<sup>2</sup>) | 0.98 (0.96–0.99) | 0.006 | 0.99 (0.97–1.00) | 0.142 |

\*Adjusted for age, sex, body mass index, hypertension, diabetic duration, proteinuria, and estimated glomerular filtration rate.
HR, hazard ratio; CI, confidence interval; PCR, random urine protein-to-creatinine ratio; eGFR, estimated glomerular filtration rate.

To eliminate the potential contribution of the urine creatinine denominator to the outcome, a sensitivity analysis was conducted using the urine abundance of proteins without normalization for urine creatinine. These analyses yielded the same results, with cluster 2 and the high complement score group both significantly associated with rapid DKD progression, compared to cluster 1 and the low complement score group, respectively (Supplementary Figure 3).

### Histopathologic correlation with complement scores in SNUH-DN cohort

In the SNUH-DN cohort, which included paired urine and kidney tissues from the same individuals, urine complement scores were significantly correlated with three of five features used to classify DKD: glomerular lesions, interstitial fibrosis and tubular atrophy (IFTA), and arteriolar hyalinosis. The correlation between complement score and interstitial inflammation did not reach statistical significance. Urine complement score was not correlated with arteriosclerosis in this sample set (Supplementary Figure 4).

### Baseline characteristics in the CRIC-T2D cohort

The association between urinary complement proteins and kidney disease progression was examined in an independent cohort: the CRIC-T2D. The CRIC-T2D cohort (Table 4), comprising 282 patients, was a majority (62%) male group with a mean age of 60.8 ± 7.5 years. Of these patients, 51.8% were non-Hispanic black and 30.1% were non-Hispanic white. Hypertension and cardiovascular disease were identified in 93.6% and 46.5% of the participants, respectively. The median 24-hour urine protein and eGFR were 0.48 g (IQR, 0.10–1.87) and 42 mL/min/1.73 m² (IQR, 37–49), respectively. The median follow-up duration was 9.7 years (IQR, 6.3–12.0).

**Table 4.** Baseline characteristics of the CRIC-T2D cohort.

| Variables | Total<br>(n = 282) | No progression<br>(n = 101) | Progression<br>(n = 181) | *P* |
| --- | --- | --- | --- | --- |
| Age (years) | 60.8 ± 7.5 | 61.8 ± 7.3 | 60.3 ± 7.5 | 0.103 |
| Male (%) | 62.4 | 58.4 | 64.6 | 0.365 |
| Body mass index (kg/m<sup>2</sup>) | 34.7 ± 7.6 | 34.7 ± 8.2 | 34.7 ± 7.2 | 0.997 |
| Race and ethnicity |  |  |  |  |
| Other | 18.1 | 11.9 | 21.5 | 0.105 |
| White non-Hispanic | 30.1 | 34.7 | 27.6 |  |
| Black non-Hispanic | 51.8 | 53.5 | 50.8 |  |
| Hypertension (%) | 93.6 | 91.1 | 95.0 | 0.297 |
| Cardiovascular disease (%) | 46.5 | 44.6 | 47.5 | 0.724 |
| Hemoglobin A1c (%) | 7.1 (6.3–8.1) | 7.0 (6.2–7.8) | 7.1 (6.4–8.2) | 0.128 |
| 24-hour urine protein (g) | 0.48 (0.10–1.87) | 0.10 (0.05–0.38) | 1.03 (0.30–3.30) | <0.001 |
| eGFR (mL/min/1.73 m<sup>2</sup>) | 42 (37–49) | 43.7 (38–48) | 41 (35–49) | 0.031 |
| Follow-up time (years) | 9.7 (6.3–12.0) | 9.7 (5.2–12.0) | 9.7 (6.5–12.0) | 0.668 |

eGFR, estimated glomerular filtration rate.

### Complement Score in the CRIC-T2D cohort

In the CRIC-T2D cohort, targeted urinary proteomics was used to quantify 12 complement proteins, including complement activators such as factor B (CFB), C3, component 4 (C4), C5, C6, component 7 (C7), component 8, alpha chain (C8A), and component 9 (C9), and inhibitors such as CD59, CFI, clusterin, and CFH-related protein 2 (Supplementary Table 2 and Supplementary Figure 5). The complement score was calculated for each participant as described in the Complement scores section. The median complement score for all participants was 1.07 (IQR, 0.42–2.10), ranging from –0.76 to 3.04. Based on this median value, the participants were divided into groups with high (>1.07) and low (≤1.07) complement scores (Figure 4A). The median complement scores for the high and low groups were 2.16 (IQR, 1.63–2.45) and 0.41 (IQR, 0.17–0.68), respectively. The group with high scores had a 2.51-fold (95% CI, 1.76–3.57) higher risk of kidney disease progression than the low score group, after adjustment for age, sex, body mass index, hypertension, ethnicity and race, 24-hour urine protein, and eGFR (Figure 4B and Table 5). Multivariable-adjusted spline regression revealed a linear association between the complement score and the hazard of DKD progression (Figure 4C).

**Figure 4.**
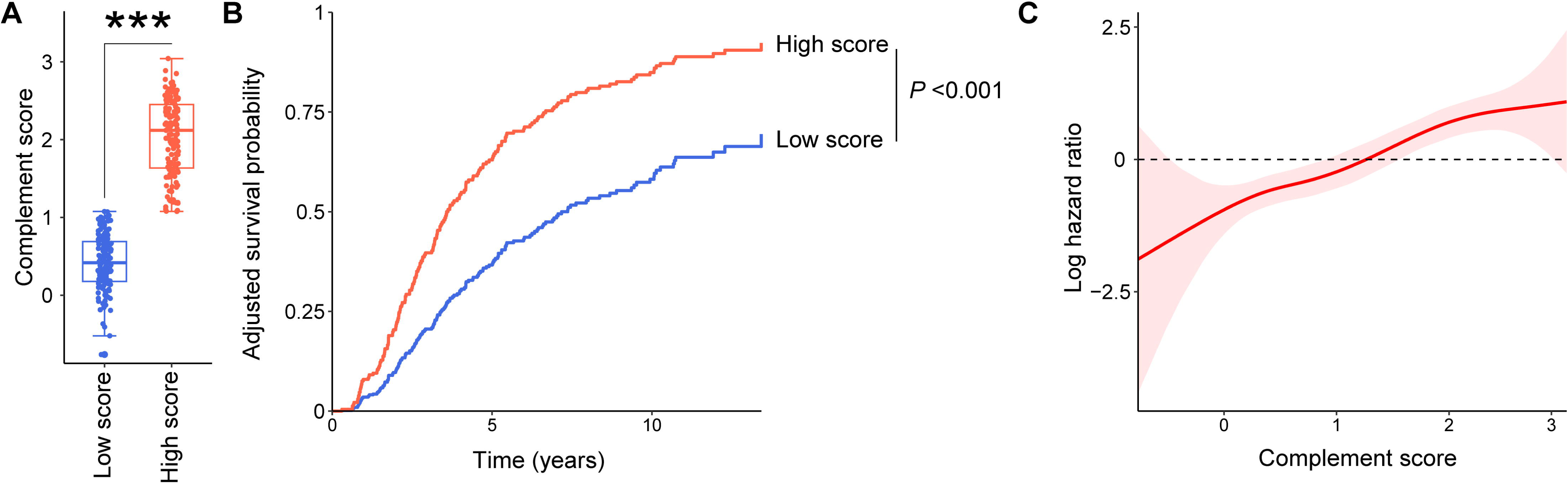
Complement score and kidney disease progression in the CRIC-T2D cohort. **A.** Bar plot of complement score in high and low score groups. \*\*\**P* <0.001. **B.** Multivariable-adjusted cumulative incidence of kidney disease progression according to complement score groups after adjustment for age, sex, body mass index, race, hypertension, 24-hour urine protein, and eGFR. **C.** Penalized spline regression according to complement score after adjusting for age, sex, body mass index, race, hypertension, 24-hour urine protein, and eGFR.

**Table 5.** Association between complement score and kidney disease progression in the CRIC-T2D cohort.

| Variables | Univariable |  | Multivariable* |  |
| --- | --- | --- | --- | --- |
|  | HR (95% CI) | *P* | HR (95% CI) | *P* |
| High score (vs. low) | 3.76 (2.74–5.16) | <0.001 | 2.51 (1.76–3.57) | <0.001 |
| Age (per 1 year) | 0.98 (0.96–1.00) | 0.120 | 1.00 (0.97–1.02) | 0.720 |
| Female (vs. male) | 1.02 (0.75–1.38) | 0.903 | 1.21 (0.87–1.68) | 0.263 |
| Body mass index (per 1 kg/m<sup>2</sup>) | 1.00 (0.98–1.02) | 0.682 | 1.00 (0.97–1.02) | 0.822 |
| Race and ethnicity (vs. other) |  |  |  |  |
| White non-Hispanic | 0.55 (0.36–0.49) | 0.707 | 0.79 (0.51–1.23) | 0.296 |
| Black non-Hispanic | 0.01 (0.84–1.03) | 0.070 | 0.96 (0.65–1.42) | 0.839 |
| Hypertension (vs. none) | 1.60 (0.82–3.13) | 0.171 | 1.71 (0.87–3.37) | 0.121 |
| 24-hour urine protein (per 1 g) | 1.31 (1.25–1.37) | <0.001 | 1.23 (1.17–1.30) | <0.001 |
| eGFR (per 1 mL/min/1.73 m<sup>2</sup>) | 0.97 (0.96–0.99) | 0.003 | 0.99 (0.97–1.01) | 0.263 |

\*Adjusted for age, sex, body mass index, race and ethnicity, hypertension, 24-hour proteinuria, and estimated glomerular filtration rate.
HR, hazard ratio; CI, confidence interval; eGFR, estimated glomerular filtration rate.

## Discussion

We used untargeted and targeted mass spectrometry to investigate the association between complement proteins and DKD progression in the SNUH-DN and CRIC-T2D cohorts. Unsupervised clustering of proteins identified in untargeted urinary proteomics from the SNUH-DN cohort revealed two groups with different kidney outcomes. Pathway analysis revealed a marked prominence of complement proteins in the group with rapidly progressive DKD. Complement scores, constructed based on the abundance and function of complement proteins, were associated with histopathologic severity and pace of DKD progression. These results were replicated in the CRIC-T2D cohort, where high complement scores similarly correlated with rapid progression. Together, these findings underscore the potential of urinary complement proteins as predictive biomarkers for kidney disease progression and the pathway as a target for therapeutic intervention in DKD.

While agents, including renin-angiotensin-system inhibitors, sodium-glucose cotransporter-2 inhibitors, and mineralocorticoid receptor antagonists, have improved kidney outcomes in DKD patients, a significant number still experience rapid disease progression ^44, 45^. The underlying pathophysiology contributing to this residual risk may involve the complement pathway, characterized by an imbalance between activators and inhibitors ^46, 47^. High urine levels of complement proteins in DKD, comparable to those in autoimmune glomerulonephritis, have been associated with worse kidney outcomes ^48, 49^. Even in patients with biopsy-confirmed DN, the association of complement proteins with worse outcomes has been observed ^50, 51^. Our results support and extend these findings and highlight the importance of further exploring the involvement of the complement pathway in DKD progression.

We add strengths to prior studies in several ways. Firstly, we combined the use of untargeted and targeted proteomics. Employing untargeted proteomics enabled an unbiased examination of the urinary proteome, leading to the initial identification of an increase in complement pathway proteins. Targeted proteomics was then utilized to validate these observations. Secondly, to our knowledge, this is the first study to replicate the association between urine complement components and DKD progression in two independent cohorts. Thirdly, our approach goes beyond examining individual complement proteins and measures a complement score, which accounts for both activator and inhibitor functions, similar to methodologies in previous studies that used expression levels of molecules involved in specific pathways for scoring ^36, 37^. This approach allowed both a qualitative and quantitative analysis by categorizing patients into high and low complement score groups and using spline regression to analyze the shape of the association between the complement score and the hazard of disease progression.

We identified several complement activators, such as CFD, C3, C4, C5, C6, C8, and mannan-binding lectin serine protease 1 and 2, that were elevated in patients with rapid kidney disease progression. Upregulation of several complement proteins (e.g., C3, component 5 alpha chain (C5A), mannan-binding lectin serine protease 1 and 2) has been noted in rats receiving a high-fat diet combined with low-dose streptozotocin ^52^. Previous human data on DKD identified C4 and C8 as significant risk factors for ESKD and all-cause mortality ^49^. Inhibiting these activators may serve as a therapeutic intervention, as one previous study showed that an inhibitor targeting the C5A receptor attenuated kidney fibrosis in streptozotocin-treated mice ^53^. The monoclonal antibody targeting C5, eculizumab, is an established clinical intervention in atypical hemolytic uremic syndrome and also tried in glomerulonephritis such as immunoglobulin A nephropathy and anti-neutrophil cytoplasmic antibody-mediated vasculitis ^54, 55^. As a C3 inhibitor, pegcetacoplan, originally effective for paroxysmal nocturnal hematuria, is currently undergoing clinical trials as a treatment for C3 glomerulopathy and immune complex-mediated membranoproliferative glomerulonephritis ^56, 57^. The time is approaching to examine complement inhibition as a novel treatment in DKD.

We observed an increase in some complement inhibitors (e.g., CFH, CLU, and CFI) and a drop in others (CD55, CD59) in rapid progressors. These findings are consistent with previous studies, which have noted similar patterns of increase in other inhibitors alongside decreased CD55 during DKD progression ^51^. The increased complement inhibitors may suggest a compensatory response to an activated complement pathway, perhaps displaying an inadequate regulatory mechanism that is insufficient to counterbalance the activation of the complement pathway ^49^. CD55 depletion exacerbates DKD in an animal model.^58^ Low urine CD59 was associated with an increased risk of ESKD, cardiovascular complications, and all-cause mortality in patients with DKD^49, 59^, and overexpression of CD55 and CD59 attenuated kidney damage in ischemia-reperfusion injury mouse models ^60^. Accordingly, CD55 and CD59 may exhibit a different pattern and thus play different roles in complement regulation compared to other complement inhibitors, which showed a compensatory increase in DKD.

Although the study provides valuable insights, there are certain limitations that warrant further examination. As with other human studies, except randomized controlled trials, it is challenging to confirm causality and elucidate the underlying mechanisms solely based on our findings. We did not examine the trend of changes in complement proteins over time, which limits our understanding of their dynamics. Additionally, we did not identify the source of the complement proteins. While the liver is considered a major source of complement proteins, the kidney may also contribute significantly, especially in the context of injury and inflammation ^36, 61^. Furthermore, the present cohorts primarily consisted of patients with reduced eGFR or advanced stage of DKD, hindering the application of the results to those with earlier stages of DKD.

We find a significant association between the complement pathway and rapid DKD progression in two independent populations. Our findings highlight urine complement proteins as potential biomarkers of pathway status and the complement pathway itself as a potential therapeutic target. Comparable findings in two ethnically dissimilar cohorts suggest that current observations are likely generalizable. By integrating various complement proteins and validating the results in an independent cohort, we provide robust evidence for the involvement of the complement pathway in rapidly progressive DKD. These findings lay the foundation for its clinical application and advocate for continued research into complement-targeting interventions to improve outcomes in the complex landscape of DKD.

## Data Availability

All data produced in the present study are available upon reasonable request to the authors.

https://www.proteomexchange.org/

## Acknowledgment

Not applicable.

## Authors’ contributions

SSH and MA designed the study. DY, SB analyzed the collected data. YG, CDN, TLF, WJQ performed targeted proteomics. LL analyzed the targeted proteomic data. DH performed untargeted proteomics. KCM reviewed kidney biopsy section and classified histopathologic parameters. SSH, DKK and YSK assisted the development of the SNUH-DN cohort and sample collections. MA was the recipient of an NIH award with access to CRIC-T2D samples. MA and WW supervised and funded the targeted proteomics study in CRIC-T2D. AZR and PS assisted the interpretation of untargeted proteomics and assisted the evaluation of histopathologic parameters. DY, SSH, and MA wrote the manuscript. SSH and MA critically reviewed the manuscript. All authors approved the final manuscript.

## Availability of data and materials

The datasets generated during and/or analyzed during the current study are available from the corresponding author upon reasonable request. The untargeted proteomic data have been deposited in the ProteomeXchange Consortium (http://proteomecentral.proteomexchange.org) via the PRIDE partner repository (accession no. PXD037505).

