## Supplementary Figure 1 for "Urine complement proteins are associated with kidney disease progression of type 2 diabetes in Korean and American cohorts"

**Supplementary Figure 1.** Heatmap of intensity-based absolute quantifications for proteins in the SNUH-DN cohort with Bonferroni adjustment. **A.** Abundant proteins in cluster 2 compared to cluster 1. **B.** Scarce proteins in cluster 2 compared to cluster 1.

**A**

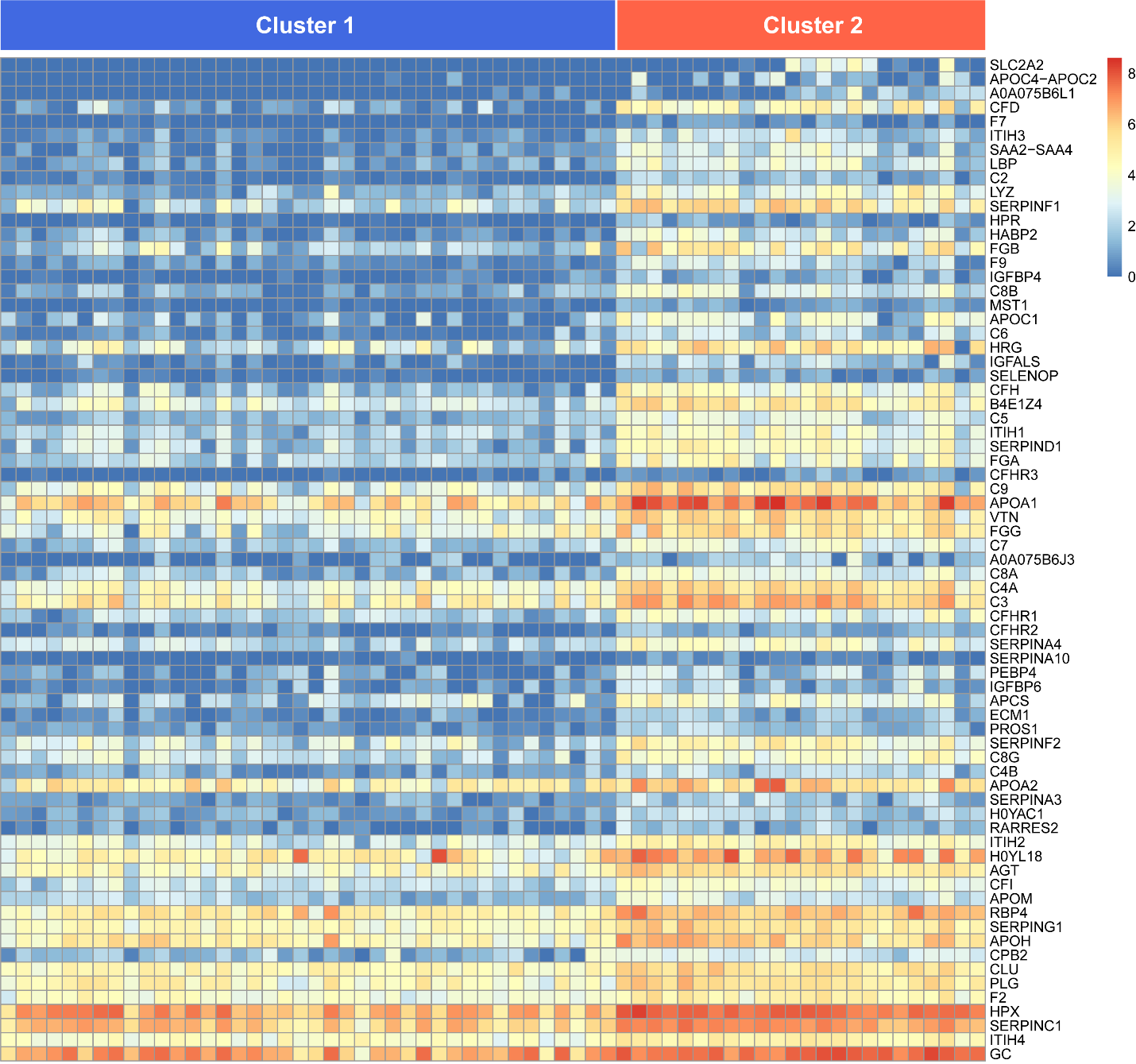

**B**

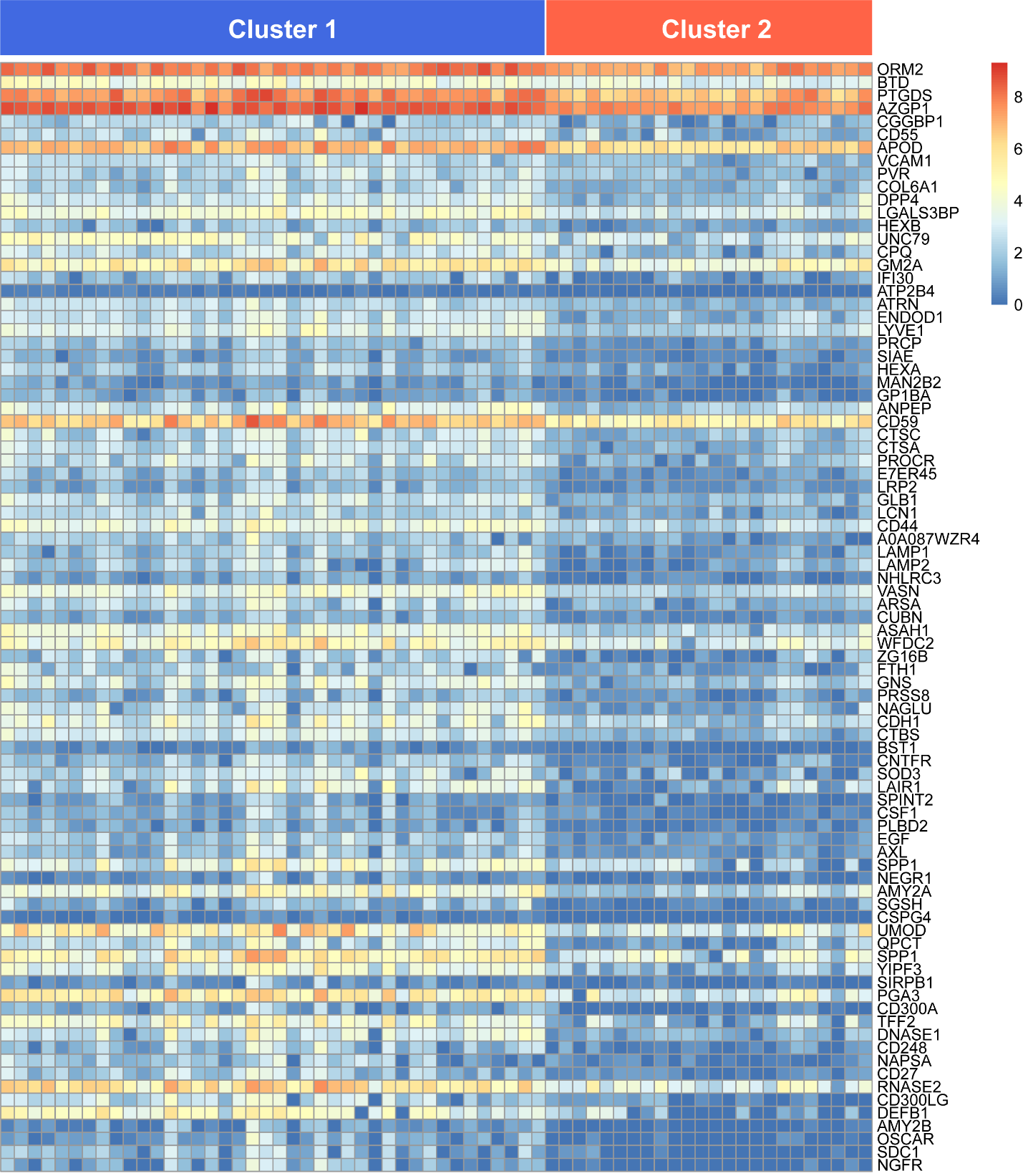
