## Supplementary Figure 2 for "Urine complement proteins are associated with kidney disease progression of type 2 diabetes in Korean and American cohorts"

**Supplementary Figure 2.** Complement scores in the SNUH-DN cohort. Patients were binarized into high and low groups according to the median complement score. ****P* <0.001.

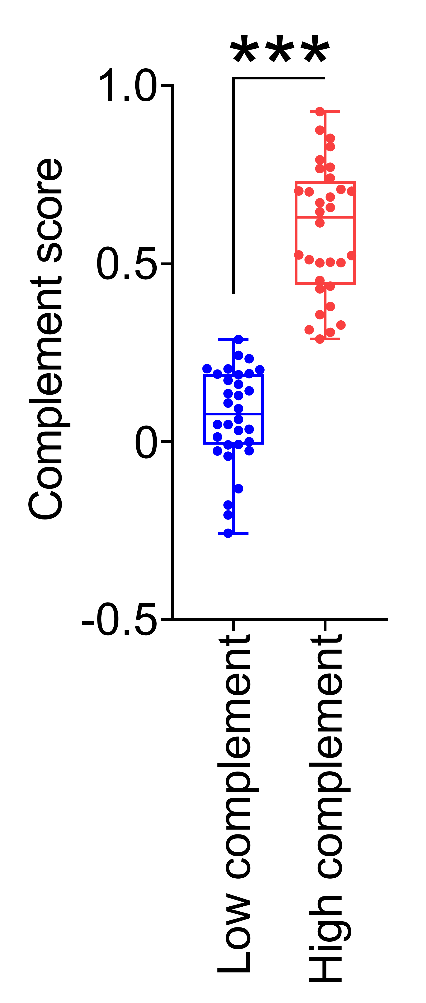
