## Supplementary Figure 3 for "Urine complement proteins are associated with kidney disease progression of type 2 diabetes in Korean and American cohorts"

**Supplementary Figure 3.** Rates of kidney disease progression using proteins not normalized by urine creatinine in the SNUH-DN cohort, after adjusting for age, sex, body mass index, hypertension, diabetic duration, proteinuria, and estimated glomerular filtration rate. **A.** Comparison between clusters 1 and 2. **B.** Comparison between high and low complement score groups.

**A**	**B**

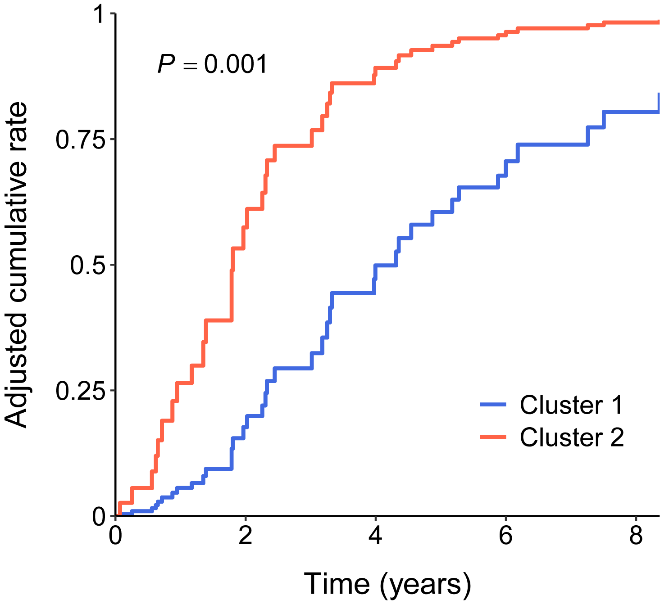

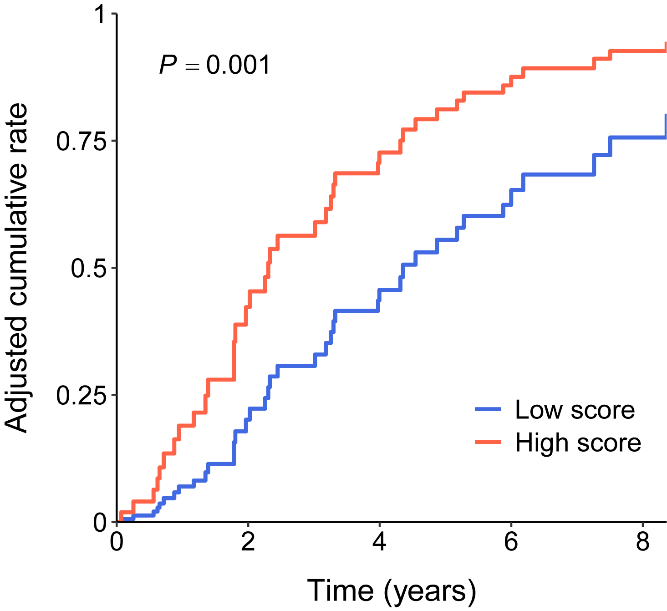
