## Supplementary Figure 4 for "Urine complement proteins are associated with kidney disease progression of type 2 diabetes in Korean and American cohorts"

**Supplementary Figure 4.** Correlation between complement scores and histopathologic parameters in the SNUH-DN cohort. Asterisk next to each lesion represents significance for trend. **P* <0.05; ***P* <0.01. IFTA, interstitial fibrosis and tubular atrophy.

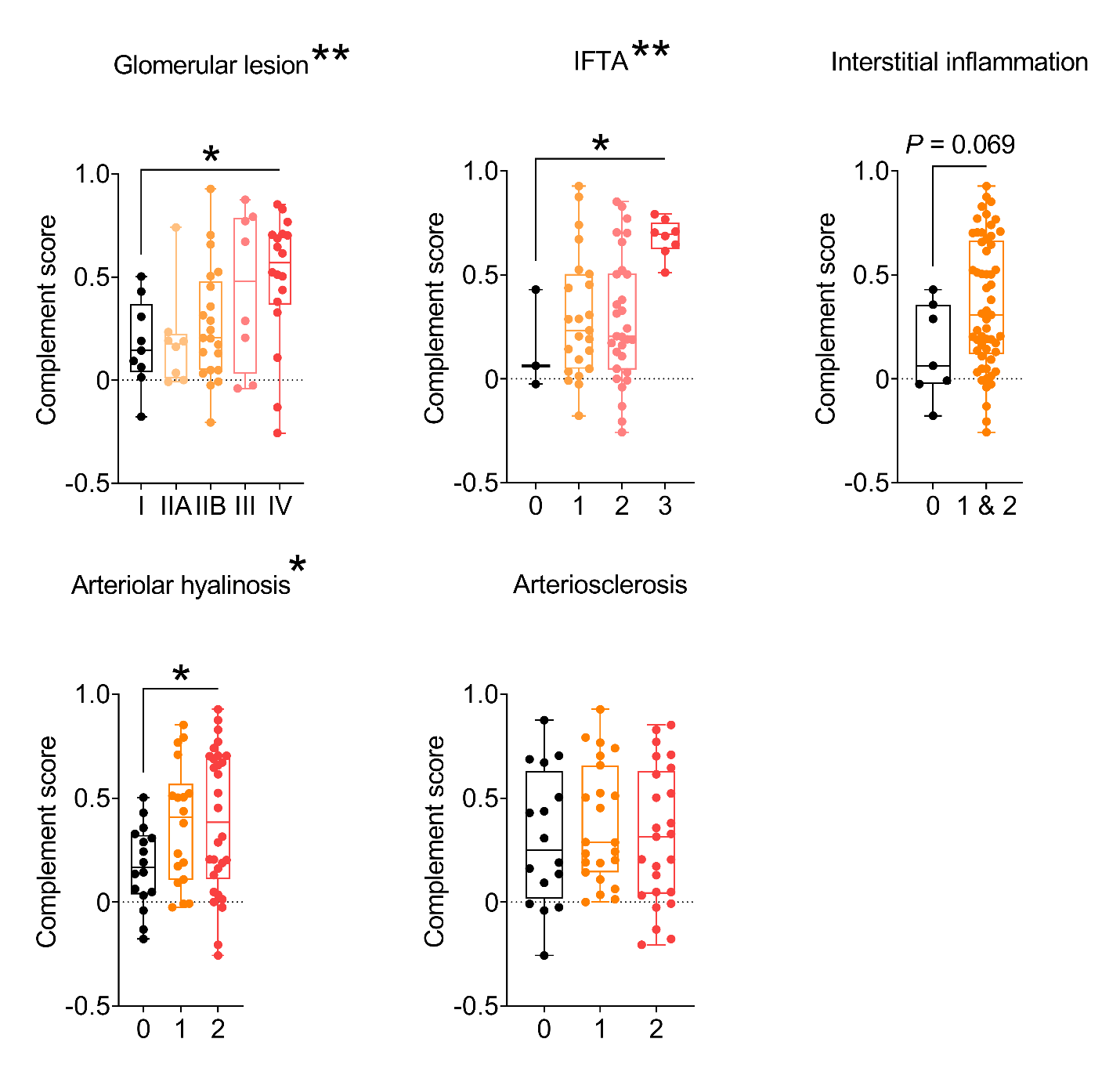
