## Supplementary Figure 5 for "Urine complement proteins are associated with kidney disease progression of type 2 diabetes in Korean and American cohorts"

**Supplementary Figure 5.** Heatmap of protein abundance for urinary complement components according to complement score groups in the CRIC-T2D cohort.

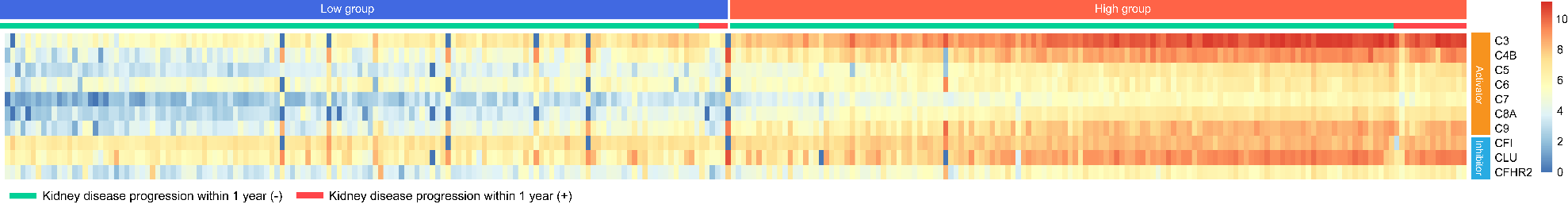
