## Supplementary Table 1 for "Urine complement proteins are associated with kidney disease progression of type 2 diabetes in Korean and American cohorts"

**Supplementary Table 1.** Peptide sequences of targeted proteomics used in LC-MS

| Protein Name | Gene | UniProt ID | Peptide Sequences | | |
| --- | --- | --- | --- | --- | --- |
| CD59 glycoprotein | *CD59* | P13987 | AGLQVYNK | ENELTYYCCK | FEHCNFNDVTTR |
| Clusterin | *CLU* | P10909 | ELDESLQVAER | IDSLLENDR |  |
| Complement 3 | *C3* | P01024 | VHQYFNVELIQPGAVK | VLLDGVQNPR | VYAYYNLEESCTR |
| Complement 4B | *C4B* | P0C0L5 | GLCVATPVQLR | LTVAAPPSGGPGFLSIERPDSRPPR | VTASDPLDTLGSEGALSPGGVASLLR |
| Complement 5 | *C5* | P01031 | ALVEGVDQLFTDYQIK | TDAPDLPEENQAR |  |
| Complement 6 | *C6* | P13671 | GEVLDNSFTGGICK | IGESIELTCPK | YYQENFCEQICSK |
| Complement 7 | *C7* | P10643 | ELSHLPSLYDYSAYR | LIDQYGTHYLQSGSLGGEYR | VLFYVDSEK |
| Complement 8 alpha chain | *C8A* | P07357 | AIDEDCSQYEPIPGSQK | LGSLGAACEQTQTEGAK | YNPVVIDFEMQPIHEVLR |
| Complement 8 beta chain | *C8B* | P07358 | IPGIFELGISSQSDR | LPLEYSYGEYR | SGFSFGFK |
| Complement 8 gamma chain | *C8G* | P07360 | RPASPISTIQPK | SLPVSDSVLSGFEQR | VQEAHLTEDQIFYFPK |
| Complement 9 | *C9* | P02748 | AIEDYINEFSVR | LSPIYNLVPVK | TSNFNAAISLK |
| Complement factor B | *CFB* | P00751 | EELLPAQDIK | VSEADSSNADWVTK |  |
| Complement factor D | *CFD* | P00746 | ATLGPAVRPLPWQR | RPDSLQHVLLPVLDR |  |
| Complement factor H-related protein 2 | *FHR2* | P36980 | LVYPSCEEK | TGDIVEFVCK |  |
| Complement factor I | *CFI* | P05156 | GLETSLAECTFTK | VFCQPWQR |  |
