## Supplementary Table 2 for "Urine complement proteins are associated with kidney disease progression of type 2 diabetes in Korean and American cohorts"

**Supplementary Table 2.** List on complement proteins. Intensity of the components was used to calculate the cumulative complement score

| Accession | Gene | Description | Action |
| --- | --- | --- | --- |
| D6R934 | *C1QB* | Complement C1Q, subcomponent B chain | Activator |
| P02747 | *C1QC* | Complement C1Q, subcomponent C chain | Activator |
| F5H2D0 | *C1R* | Complement C1, subcomponent R | Activator |
| Q9NZP8 | *C1RL* | Complement C1, R subcomponent-like | Activator |
| P06681 | *C2* | Complement C2 (C3/C5 convertase) | Activator |
| P01024 | *C3* | Complement C3 | Activator |
| P0C0L4 | *C4A* | Complement C4, isotype A | Activator |
| P0C0L5 | *C4B* | Complement C4, isotype B | Activator |
| P01031 | *C5* | Complement C5 | Activator |
| P13671 | *C6* | Complement C6 | Activator |
| P10643 | *C7* | Complement C7 | Activator |
| P07357 | *C8A* | Complement C8, alpha chain | Activator |
| P07358 | *C8B* | Complement C8, beta chain | Activator |
| P07360 | *C8G* | Complement C8, gamma chain | Activator |
| P02748 | *C9* | Complement C9 | Activator |
| B1AP13 | *CD55* | Decay-accelerating factor | Inhibitor |
| E9PNW4 | *CD59* | CD59 glycoprotein | Inhibitor |
| Q9NPY3 | *CD93* | Complement C1q receptor | Activator |
| K7ERG9 | *CFD* | Complement factor D | Activator |
| P08603 | *CFH* | Complement factor H | Inhibitor |
| Q03591 | *CFHR1* | Complement factor H–related protein 1 | Inhibitor |
| P36980-2 | *CFHR2* | Complement factor H–related protein 2 (isoform 2) | Inhibitor |
| Q02985-2 | *CFHR3* | Complement factor H–related protein 3 (isoform 2) | Inhibitor |
| Q92496-2 | *CFHR4* | Complement factor H–related protein 4 (isoform 2) | Inhibitor |
| Q9BXR6 | *CFHR5* | Complement factor H–related protein 5 | Inhibitor |
| G3XAM2 | *CFI* | Complement factor I | Inhibitor |
| E9PAQ1 | *CFP* | Complement factor properdin | Activator |
| P10909-4 | *CLU* | Clusterin (isoform 4) | Inhibitor |
| P48740-4 | *MASP1* | Mannan–binding lectin serine protease 1 (isoform 4) | Activator |
| O00187-2 | *MASP2* | Mannan–binding lectin serine protease 2 (isoform 4) | Activator |
| P05155 | *SERPING1* | Plasma protease C1 inhibitor | Inhibitor |
