## Supplementary Table 3 for "Urine complement proteins are associated with kidney disease progression of type 2 diabetes in Korean and American cohorts"

**Supplementary Table 3.** Proteins upregulated in cluster 2 mapped to ten pathways

| Rank | Pathway | Relevant proteins |
| --- | --- | --- |
| 1 | Complement system | CFD, C3, C4A, C7, C8A, C5, VTN, CFH, C6, C9, C2, CFI, SERPING1, APOA1, FGG, CFHR2, FGB, PROS1, PLG, FGA, APCS |
| 2 | Complement and coagulation cascades | CFD, C3, C7, SERPIND1, CFH, C6, C8G, C9, C2, CFI, SERPING1, FGB, SERPINC1, CLU, PROS1, PLG, CPB2, F7, SERPINF2, F2, F9 |
| 3 | Network map of SARS-CoV-2 signaling pathway | LBP, ITIH3, AGT, C8A, CFH, CFI, HRG, APOA1, FGG, ITIH4, FGB, APOC1, FGA, APOH, APOM, SERPINA10, APOA2 |
| 4 | Complement system in neuronal development and plasticity | CFD, C3, C4A, C7, C8A, C8B, C5, VTN, CFH, C6, C8G, C9, C2, CFI, SERPING1, CLU, PROS1 |
| 5 | Complement activation | CFD, C3, C4A, C7, C8A, C8B, C5, C6, C8G, C9, C2 |
| 6 | Selenium micronutrient network | APOA1, FGG, FGB, PLG, FGA, F7, F2, SELENOP, SERPINA3 |
| 7 | Allograft rejection | C3, C4A, C7, C8A, C8B, C5, C6, C9, C2 |
| 8 | Folate metabolism | APOA1, FGG, FGB, PLG, FGA, F7, F2, SERPINA3 |
| 9 | Blood clotting cascade | FGG, FGB, PLG, FGA, F7, SERPINF2, F2, F9 |
| 10 | Vitamin B12 metabolism | APOA1, PLG, F7, F2, SERPINA3 |
